# CDC123 variants associated with developmental abnormalities alter CDC123-eIF2 subunit interactions and impair protein synthesis

**DOI:** 10.64898/2026.11.10.26367377

**Authors:** Anthony L. Erb, Alexandra Misciagna, Ambar Rodriguez-Martinez, Brooklynn Gasser, Colette DeFilippo, Annika E. Gillam, Daniah Beleford, Grace E. VanNoy, Sara K. Young-Baird

## Abstract

The translation initiation factor eIF2 is an essential regulator of cellular protein synthesis with direct links to human physiology and development. Composed of eIF2α, β, and γ subunits, eIF2 heterotrimer assembly requires the activity of its dedicated chaperone and the essential protein, CDC123. Accordingly, CDC123 is a key determinant of protein synthesis and cell growth. Previous research has demonstrated that variants in eIF2γ impair neurodevelopment, and *CDC123* variants of uncertain significance have recently been reported for pediatric patients with neurologic findings. However, the potential pathogenicity, and cell and molecular impacts of these clinically relevant *CDC123* variants remains entirely unexplored. In this work, we describe four unrelated families with pediatric patients that present with neurodevelopmental delays, hypotonia, and motor dysfunction. The probands are heterozygous for three novel CDC123 missense variants. The potential pathogenicity of the variants is comprehensively assessed through appraisal of *CDC123* variant allele frequencies, protein conservational analyses, structural modeling, and functional assays in both humanized yeast and human cell-based model systems. Cellular and biochemical studies indicated that the variants are damaging to CDC123 activity, altering CDC123-eIF2 subunit interactions and leading to reduced cell growth. Furthermore, we demonstrate that the *CDC123* variants display dominant-negative effects on yeast cell growth and protein synthesis, rather than causing haploinsufficiency. Together, our findings provide the first link between *CDC123* missense variants and human disease and highlight that CDC123-mediated eIF2 heterotrimer formation is essential for human development.

## INTRODUCTION

The regulation of mRNA translation fundamentally shapes gene expression, and both cellular and human physiology. For instance, mRNA translational control underlies how cells respond to internal and external stressors, including amino acid deprivation, viral infection, and UV irradiation, and drives critical cell fate “decisions” to either adapt to the stress or undergo cell death^1–4^. Deficits in this process and pathogenic variants in the translational machinery invariably result in human diseases, many of which have pressing knowledge gaps and lack effective treatment options^5,6^.

The Integrated Stress Response (ISR) is a centralized mechanism by which cells modulate protein synthesis under various stress conditions. During canonical activation of the ISR, specialized stress-sensing kinases phosphorylate the α subunit of the translation initiation factor 2 (eIF2)^2,7^. eIF2 is composed of three essential protein subunits, eIF2α, β, and γ, and facilitates translation initiation by delivering the initiator methionyl tRNA to the ribosome^8^. Phosphorylation of eIF2α (eIF2α-P) reduces eIF2 activity, resulting in a decrease in general protein synthesis and, thus, conservation of energy resources during cellular stress.

Importantly, protein synthesis levels are also regulated by formation of the eIF2 heterotrimer itself, which requires the activity of the eIF2 chaperone, cell division cycle 123 (CDC123)^9,10^. The eIF2α, β, and γ subunits and CDC123 are all essential proteins^11,12^. eIF2γ serves as the core of the eIF2 complex and makes contacts with eIF2α and eIF2β, which do not directly interact with each other^13,14^. CDC123 was previously shown to promote eIF2 complex assembly by binding eIF2α and eIF2γ to facilitate eIF2α-eIF2γ heterodimer formation, while eIF2β binds to eIF2γ in a CDC123-independent manner^9,15,16^.

Variants in *EIF2S3*, the gene encoding eIF2γ, cause MEHMO syndrome, an X-linked intellectual disability disorder that features additional motor and developmental delays^15,17–28^. In our previous studies of MEHMO syndrome, we found that a specific disease-causing variant at the eIF2γ C-terminus altered the eIF2γ-CDC123 interaction site, impairing the ability of CDC123 to promote eIF2 complex formation and reducing overall levels of protein synthesis and cell growth^15^. This prompted us to explore variants in genes related to *EIF2S3* and, in this report, to assess recently identified variants of uncertain significance in the autosomal gene *CDC123* that are found in patients that presented with clinical features shared with MEHMO syndrome.

Here, we describe four probands with three, heterozygous, novel missense variants in *CDC123* who display developmental abnormalities including neurodevelopmental delays, hypotonia, and motor dysfunction. This work reports the first genetic, cellular, and biochemical assessments to suggest a link between *CDC123* missense variants and human disease. We provide familial pedigrees and clinical phenotyping, combined with structural and *in silico* pathogenicity assessments of the *CDC123* missense variants. Additionally, we functionally characterize the *CDC123* variants in both yeast and human cell model systems and determine that the variants impair CDC123 activity. We find that the variants map to different domains in CDC123 and disrupt CDC123-eIF2 subunit binding through distinct mechanisms. Altered eIF2 integrity impairs cell growth, which can be rescued by eIF2 subunit overexpression.

Furthermore, we demonstrate that the *CDC123* variants display dominant-negative effects on yeast cell growth and protein synthesis, rather than causing haploinsufficiency. Together, our results suggest these CDC123 missense variants are likely pathogenic and impair critical CDC123-eIF2 subunit interactions important for eIF2 complex formation.

## RESULTS

### Identification of variants in the autosomal gene *CDC123* in pediatric patients with developmental abnormalities

### Proband 1, Family 1

The proband was a male (Figure 1A; II:1) that presented with hypotonia, muscle weakness, motor delays, hand contractures, laryngomalacia, and obstructive sleep apnea at the time of examination. Duo exome sequencing (ES) of the proband and mother identified a novel heterozygous *CDC123* variant (NC_000010.11:g.12210292G>T, NM_006023.3:c.207G>T, NP_006014.2:p.(Trp69Cys or W69C)) in the proband that was not maternally inherited (Figure 1A, Supplemental Figure 1A and 1B, and Table 1). Sequencing data is not available for the unaffected father.

**Figure 1.**
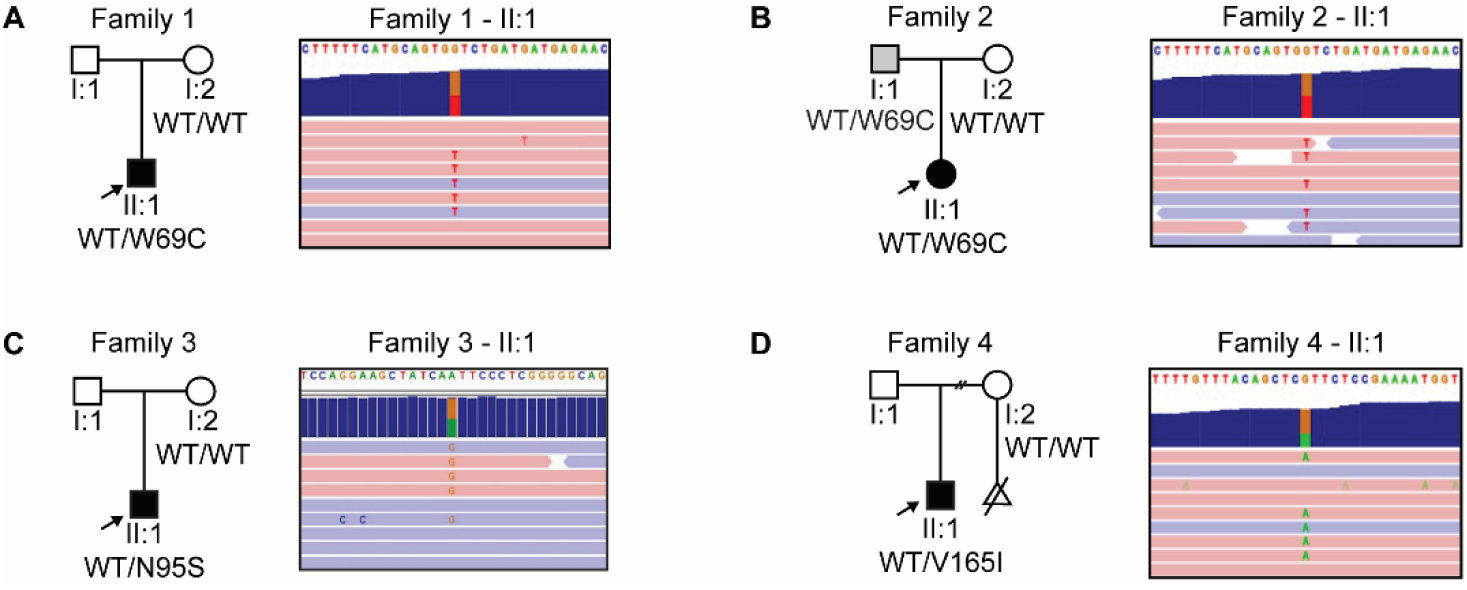
*CDC123* variants identified by exome sequencing are linked to developmental abnormalities. Familial pedigrees and identification of heterozygous *CDC123* missense variants **(A and B)** c.207G>T (p.W69C), **(C)** c.284A>G (p.N95S), and **(D)** c.493G>A (p.V165I). Probands are shown as filled symbols and are also indicated by an arrow in the familial pedigrees. Additional symbols denote the following: open square = unaffected male, open circle = unaffected female, shaded square (grey) = male *CDC123* variant carrier whose medical history includes neurologic findings. Exome sequencing results for the probands are depicted using the Integrative Genomics Viewer (IGV). Blue and red bars indicate sequencing reads from the positive (blue) and negative (red) DNA strands. *CDC123* variant sequencing reads display a red “T” (c.207G>T, p.W69C), orange “G” (c.284A>G, p.N95S), or green “A” (c.493G>A, p.V165I) in the middle of the blue and red bars. The WT *CDC123* sequence is shown at the top of the IGV image.

**Table 1.** Overview of the clinical features of affected patients with *CDC123* variants. The proband number (from the Results), variant, age at examination, inheritance, clinical descriptions, and ACMG classification of the probands are shown. N/A indicates that the data is not available.

| **Proband** | **Protein Variant (NP_006014.2)** | **Age at examination** | **Inheritance** | **Developmental delay** | **Behavioral difficulties** | **Motor and muscle findings** | **ACMG Classification** |
|---|---|---|---|---|---|---|---|
| 1 | p.(Trp69Cys) | <10 | Not maternal;<br>Unknown if paternal | Moderate | N/A | Motor delay, hypotonia, muscle weakness, hand contractures, laryngomalacia | Likely Pathogenic (PS3, PM1, PM2, PP3) |
| 2 | p.(Trp69Cys) | <10 | Paternal | Moderate | Yes (autism and ADHD) | N/A | Likely Pathogenic (PS3, PM1, PM2, PP3) |
| 3 | p.(Asn95Ser) | <10 | Not maternal;<br>Unknown if paternal | Severe | N/A | Hypotonia, muscle spasms, spasticity, and seizures.<br>Wears foot braces. | Likely Pathogenic (PS3, PM1, PP3) |
| 4 | p.(Val165Ile) | <10 | Not maternal;<br>Unknown if paternal | Moderate | Yes (autism and ADHD) | Central and peripheral hypotonia, frequent falls, spinal asymmetry (<10 degrees) | Likely Pathogenic (PS3, PM1, PP3) |

### Proband 2, Family 2

The proband was a female (Figure 1B; II:1) that displayed global developmental delay, autism, and attention-deficit/hyperactivity disorder (ADHD) at the time of examination. ES of the proband identified the same heterozygous *CDC123* variant identified for proband 1 (NC_000010.11:g.12210292G>T, NM_006023.3:c.207G>T, NP_006014.2:p.(Trp69Cys or W69C)) (Figure 1A and 1B, and Supplemental Figure 1A and 2A). Maternal and paternal ES revealed that proband 2 inherited the *CDC123* variant from the father whose medical history includes ADHD, anxiety, and depression (Figure 1B, Supplemental Figure 2B and C, and Table 1).

**Figure 2.**
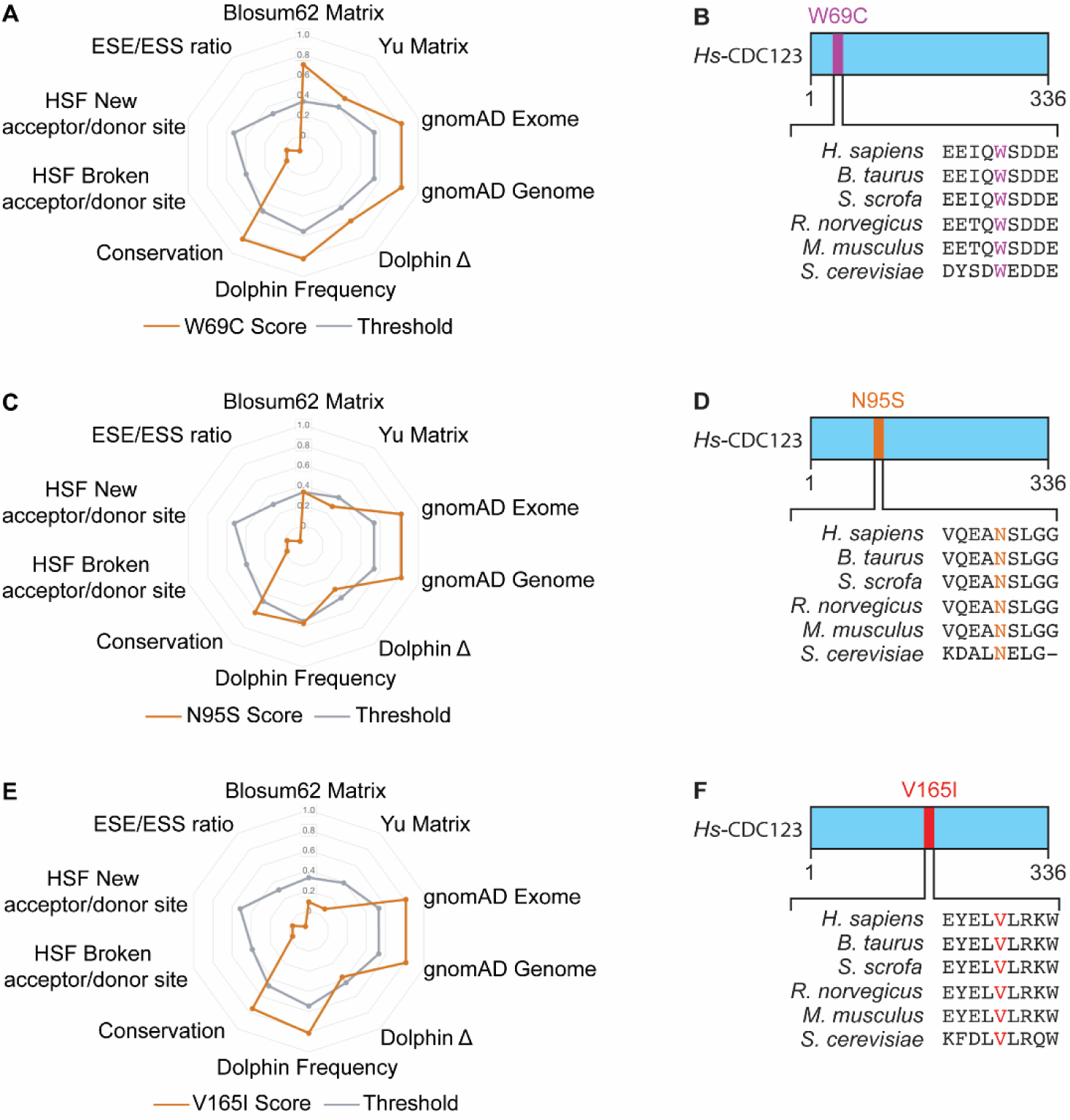
CDC123-W69C, -N95S, and -V165I are predicted to be pathogenic. UMD Predictor Pro results and CDC123 protein diagrams with amino acid alignments for the *CDC123* **(A-B)** c.207G>T (p.W69C), **(C-D)** c.284A>G (p.N95S), and **(E-F)** c.493G>A (p.V165I) variants. Each variant was assessed for predicted impacts on mRNA splicing (Human Splicing Finder acceptor/donor sites and the exonic splice enhancers/silencers (ESE/ESS) ratio), biochemical or structural impacts of amino acid substitutions on proteins (Blosum62 and Yu matrices), localization within important CDC123 protein domains (Dolphin Frequency and Dolphin Δ), amino acid conservation (Conservation), and allele frequency in the general population (gnomAD Exome and gnomAD Genome). The threshold score for pathogenicity is indicated by a grey line, and CDC123 variant scores are indicated by an orange line. Variant scores outside of the grey line are considered pathogenic by UMD Predictor Pro. The protein diagrams of human CDC123 with W69 (purple), N95 (orange), or V165 (red) include amino acid sequence alignments of the residues surrounding CDC123-W69, -N95, and -V165 in human CDC123 and its orthologs **(B, D, and F)**

### Proband 3, Family 3

The proband was a male (Figure 1C; II:1) who presented with severe developmental delay, hypotonia, daily neck and other muscle spasms, spasticity, and craniosynostosis secondary to poor brain growth at the time of examination. Proband 3 used foot braces. Clinical examination was also notable for microcephaly, strabismus, and cortical blindness. The proband presented with poorly controlled seizures, and the family history was notable for myoclonic epilepsy in the proband’s mother. Duo ES for the proband and mother identified a novel heterozygous *CDC123* variant (NC_000010.11:g.12215786A>G, NM_006023.3:c.284A>G, NP_006014.2:p.(Asn95Ser or N95S)) that was not maternally inherited (Figure 1C, Supplemental Figure 3A and 3B, and Table 1). Sequencing data is not available for the unaffected father.

**Figure 3.**
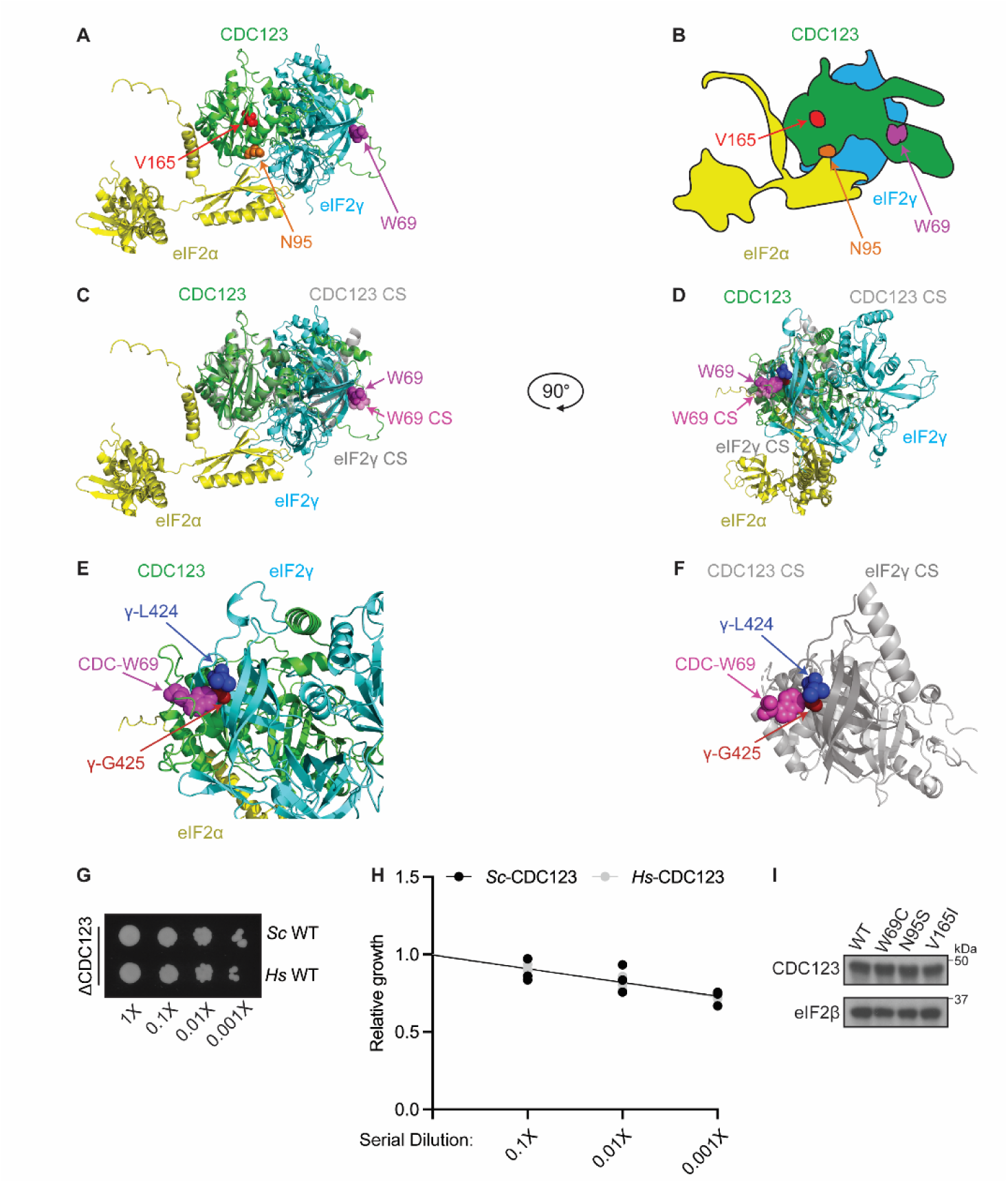
CDC123-W69, -N95, and -V165 map to distinct regions of CDC123 and do not impact protein levels in yeast models. **(A)** Ribbon representation of human CDC123 in complex with eIF2α and eIF2γ that was generated using AlphaFold2 and visualized with PyMOL Molecular Graphics System (Version 3.0 Schrödinger). Components are colored as follows: CDC123 (green), eIF2α (yellow), eIF2γ (cyan), W69 (purple), N95 (orange), and V165 (red). **(B)** Cartoon representation of the structural prediction shown in (A). Components are colored as follows: CDC123 (green), eIF2α (yellow), and eIF2γ (cyan). CDC123-W69 (purple), -N95 (orange), and -V165 (red) are shown in sphere representation. **(C)** The *Hs* CDC123-eIF2α-eIF2γ AlphaFold2 structure from 3A was overlaid with the crystal structure (CS) of *Hs* CDC123 (transparent light grey) in complex with the eIF2γ C-terminus (residues 362 to 471, transparent dark grey) that was previously reported^34^. CDC123-W69 from the CDC123 AlphaFold2 multimer (purple) and CS (transparent pink) are indicated and display a high degree of overlap. **(D)** A 90-degree rotation on the y-axis of the structure in 3C with eIF2γ-L424 (blue) and -G425 (dark red) indicated to illustrate conserved hydrophobic interactions with CDC123-W69 predicted by both the CDC123-eIF2α-eIF2γ AlphaFold2 structural model and CDC123-eIF2γ C-terminus crystal structure. **(E)** Zoomed in view of only the *Hs* CDC123-eIF2α-eIF2γ AlphaFold2 structure from 3D to better visualize the hydrophobic interactions of CDC123-W69 (purple) with eIF2γ-L424 (blue) and -G425 (dark red). **(F)** Zoomed in view of only the *Hs* CDC123-eIF2γ C-terminus crystal structure from 3D to better visualize the hydrophobic interactions of CDC123-W69 (purple) with eIF2γ-L424 (blue) and -G425 (dark red). **(G)** 10-fold serial dilutions of yeast strains expressing the WT forms of CDC123 from *Saccharomyces cerevisiae* (*Sc*) or *Homo sapiens* (*Hs*) were grown on SD minimal medium at 30°C for 3 days. **(H)** Linear regression analysis of the relative growth for biological replicate images (n=3) of the spotting assays shown in panel G and Supplemental Figure 6. The line equations and exact p-values for the comparisons discussed in panel G are reported in Supplemental Table 1. **(I)** Whole cell extracts (WCEs) from yeast strains expressing the indicated wild-type or variant forms of CDC123 were subjected to SDS-PAGE followed by immunoblotting with antibodies against human CDC123 and yeast eIF2β.

### Proband 4, Family 4

The proband was a male (Figure 1D; II:1) that displayed autism, ADHD, and had experienced two episodes of visual and auditory hallucinations by the time of examination. Clinical examination was also notable for central and peripheral hypotonia, growth deficiency and short stature (Z-score: -2.62), motor abnormalities such as frequent falls, spinal asymmetry (<10 degrees), intermittent alternating exotropia, and high astigmatism. Proband 4 is status post G-tube placement due to dysphagia and supraglottoplasty, with mild gastritis noted after endoscopy. The proband presented with hyperlipidemia (total cholesterol: 221 mg/dL, LDL: 156 mg/dL) in a setting of maternal familial hypercholesterolemia. Relevant familial history includes a maternal grandfather and uncle with records of auditory and visual hallucinations. Duo ES of the proband and mother identified a novel heterozygous *CDC123* variant (NC_000010.11:g.12235051G>A, NM_006023.3:c.493G>A, NP_006014.2:p.(Val165Ile or V165I)) that was not maternally inherited (Figure 1D, Supplemental Figure 4A, 4B and 4C, and Table 1). Sequencing data is not available for the unaffected father.

### CDC123-W69C, -N95S, and -V165I are predicted to be pathogenic variants

To first evaluate the potential pathogenicity of the CDC123-W69C, -N95S, and -V165I variants, we utilized UMD-Predictor Pro. As shown in Figure 2, this tool employs a combinatorial algorithm that takes into account anticipated impacts of single nucleotide polymorphisms on mRNA splicing (*i.e.*, Human Splicing Finder (HSF) new or broken splice acceptor/donor sites, and auxiliary exonic splicing enhancers/silencers (ESE/ESS)), predicted biochemical or structural impacts of amino acid substitutions on proteins (*i.e.*, Blosum62 Matrix, Yu Matrix, Dolphin Frequency, Dolphin Δ), amino acid conservation among species (*i.e.*, Conservation), and the prevalence of variants in the human population (*i.e.*,gnomAD Exome and gnomAD Genome)^29^. Using these criteria, UMD-Predictor Pro calculates a pathogenicity score ranging from 0-100, with any score >74 predicted to be a pathogenic variant. Notably, UMD-Predictor Pro demonstrated superior accuracy compared to other widely used pathogenicity prediction tools in an analysis of over 4,300 cancer-associated single-nucleotide variants^30^, and was thus selected for our *CDC123* variant analysis.

We found that while the *CDC123* nucleotide variants noted above were predicted to have a negligible impact on mRNA splicing signals, CDC123-W69C, -N95S, and V165I all exceeded the threshold for pathogenicity at the protein level (Figure 2A, 2C, and 2E). First, W69 is both highly conserved and located in an important CDC123 protein domain (Figure 2A; Conservation, Dolphin Frequency, and Dolphin Δ). The CDC123-W69C variant results in an R-group alteration between the WT and mutant residues and has not been previously reported in the human population (Figure 2A: Blosum62 Matrix, Yu Matrix, gnomAD Exome, and gnomAD Genome). Together these data resulted in a UMD Predictor-Pro pathogenicity score of 100/100 for CDC123-W69C. By comparison, the CDC123-N95S variant received a pathogenicity score of 82/100 primarily due to high conservation of N95 between species and low allele frequency (AF) of CDC123-N95S in the human population (AF=0.00005890 in gnomAD v4.1.0) (Figure 2C: Conservation, gnomAD Exome, and gnomAD Genome). Lastly, UMD-Predictor Pro assigned a pathogenicity score of 100/100 to the CDC123-V165I variant due to high conservation of the V165 residue, low allele frequency of CDC123-V165I in the population (AF=0.000004339 in gnomAD v4.1.0), and localization of the variant to an important CDC123 protein domain (Figure 2E: Conservation, gnomAD Exome, gnomAD Genome, and Dolphin Frequency). Thus, our *in silico* assessments predict CDC123-W69C, -N95S, and -V165I to be pathogenic.

### CDC123-W69, -N95, and -V165 are highly conserved residues that localize to distinct CDC123 protein domains

Building upon the UMD-Predictor Pro results, we conducted an independent protein conservation assessment and found that CDC123-W69, -N95, and V165 are fully conserved from yeast (*S. cerevisiae*) to humans (*H. sapiens*) (Figure 2B, 2D, and 2F). Intriguingly, the amino acid residues directly upstream and downstream of W69, N95, and V165 are also mostly conserved between species, supporting the notion that these regions of the CDC123 protein are particularly important for its function. This observation is significant as the eukaryotic CDC123 amino acid sequences assessed only share ∼30% sequence identity overall (Supplemental Figure 5).

Having confirmed that CDC123-W69, -N95, and -V165 are all highly conserved and likely functionally significant residues, we next determined their localization within the CDC123 3-dimensional protein structure. Using AlphaFold2, we generated a multimeric structural prediction of the human CDC123 protein in complex with its known interacting partners, eIF2α and eIF2γ (Figure 3A)^31^. As CDC123 specifically promotes assembly of the eIF2α-eIF2γ heterodimer and eIF2β binds to eIF2γ in a CDC123-independent fashion, eIF2β was not included in our structural model^9,16,32–34^. Importantly, the AlphaFold CDC123-eIF2α-eIF2γ structural prediction closely aligned with a recent crystal structure of human CDC123 in complex with the eIF2γ C-terminus (residues 362-471) alone (Figure 3C-3F)^34^. For instance, CDC123-W69 maps to an CDC123-eIF2γ interface (Figure 3A and 3B), and is suggested to form hydrophobic interactions with eIF2γ-L424 and -G425 in both our CDC123-eIF2α-eIF2γ model and the CDC123-eIF2γ C-terminus crystal structure (Figure 3E and 3F). In contrast to W69, CDC123-N95 localizes in close proximity to both eIF2α and eIF2γ, while V165 appears spatially remote from points of interaction with the eIF2 subunits, and more centrally located in the CDC123 protein, in the CDC123-eIF2α-eIF2γ structural model (Figure 3A and 3B). Overall, these structural insights indicate that the CDC123-W69C, -N95S, and V165I variants map to distinct regions of the CDC123 protein and likely differentially impact how CDC123 interacts with the eIF2α and eIF2γ subunits.

### CDC123 variant yeast strains display impaired cell growth that is rescued by either eIF2γ or eIF2α overexpression

To perform functional studies of the *CDC123* variants, we first generated yeast strains lacking the chromosomal *CDC123* gene and expressing plasmid-encoded wild-type yeast (*Sc* WT) or human (*Hs* WT) CDC123 as the sole source of CDC123. *CDC123* is an essential gene, so the empty vector negative control was not viable as expected.

Human CDC123 functionally complemented the yeast protein in cell growth assessments (Figure 3G, Supplemental Figure 6, quantification in Figure 3H), enabling us to directly test impacts of the human variants on CDC123 function in our yeast model system.

Introduction of CDC123-W69C, -N95S, or -V165I into our humanized yeast model did not alter CDC123 protein levels as compared to the wild-type (WT) control (Figure 3I; eIF2β was used as a protein loading control). However, the CDC123-W69C, -N95S, and -V165I yeast strains all demonstrated a marked slow growth phenotype (Figure 4A-4C: compare row 1 vs. row 4, Supplemental Figure 7, quantification in Figure 4D-4F). Building off of these results, we reasoned that if these observed growth defects were due to impaired CDC123-mediated eIF2 heterotrimer formation, then overexpression of the eIF2α and eIF2γ subunits may, through mass action, bypass the cellular requirement for CDC123 and increase eIF2 levels. Consistent with this idea and our structural models, overexpression of eIF2α and eIF2γ offered differential rescue of the slow growth phenotype for all three variants (Figure 4D-4F, and Supplemental Figure 7). Intriguingly, overexpression of eIF2γ significantly rescued the CDC123-W69C variant strain cell growth defect, consistent with the proximity of W69C to the CDC123-eIF2γ interface (Figure 3A-3F, and Figure 4A: compare row 4 vs. row 6, quantification in Figure 4D). By comparison, overexpression of eIF2α fully rescued the slow growth phenotype associated with CDC123-N95S that localizes to a CDC123-eIF2α interface. (Figure 3A, Figure 4B: compare row 4 vs. row 5, quantification in Figure 4E). Finally, the slow growth phenotype of CDC123-V165I was suppressed by overexpression of both eIF2 subunits possibly due to the location of V165 within the core structure of the CDC123 protein (Figure 3A, Figure 4C: compare rows 4 vs rows 5 and 6, quantification in Figure 4F). Together, these data indicate that the growth defects of the strains expressing the human CDC123 variants are due to impaired CDC123 function and not to altered CDC123 levels.

**Figure 4.**
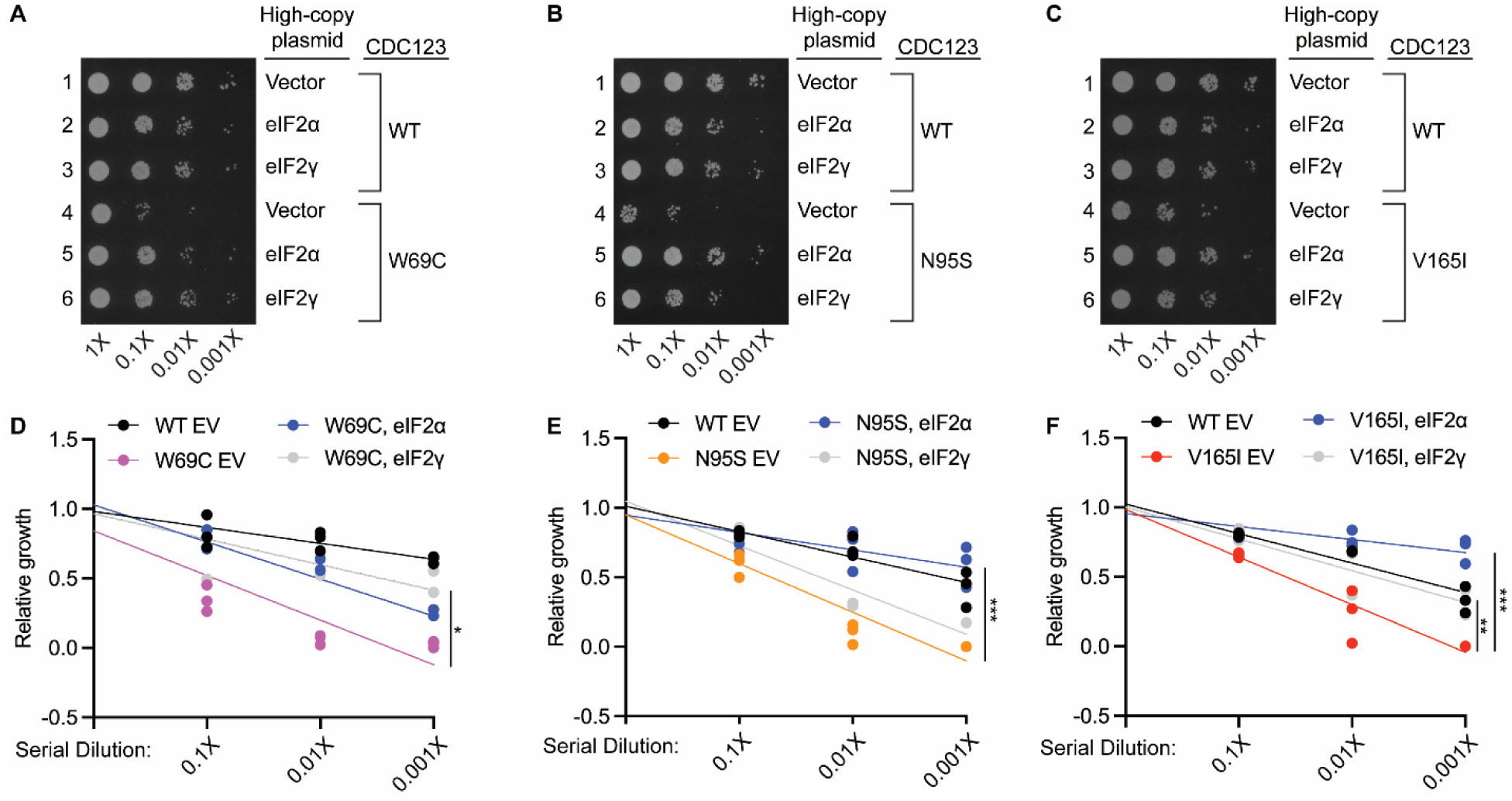
**CDC123 variants impair yeast cell growth and are differentially rescued by eIF2 subunit overexpression**. **(A-C)** 10-fold serial dilutions of yeast strains expressing the indicated WT or variant forms of human CDC123 with empty vector, or overexpression of eIF2α or eIF2γ were grown on SD minimal medium at 30°C for 3 days. **(D-F)** Linear regression analysis of the relative growth for biological replicate images (n=3) of the spotting assays shown in panels A-C. Statistically significant differences between the slopes of the indicated lines is denoted by an asterisk: *p<0.05, **p<0.005, and ***p<0.0005. The slopes and exact p-values for all comparisons discussed in panels A-C are reported in Supplemental Table 1.

### CDC123 variants cause dominant-negative effects on yeast cell growth and protein synthesis, and disrupt eIF2 subunit binding in human cells

To better recapitulate the heterozygous nature of the *CDC123* variants identified in the probands, we next introduced an additional *Hs* WT CDC123-expressing plasmid into our humanized yeast strains, generating the following CDC123 allele combinations: WT/WT, WT/W69C, WT/N95S, or WT/V165I. While the yeast strain expressing two copies of WT CDC123 grew robustly, CDC123-W69C, -N95S, and -V165I all displayed dominant-negative effects, resulting in slow growth of the CDC123 variant-containing strains (Figure 5A, 5B, and Supplemental Figure 8). Consistent with their slow growth, and the essential role of CDC123 in regulating eIF2-mediated translation initiation, all three variant-harboring strains exhibited a noticeable defect in global protein synthesis observed by decrease in the height of the 80S monosome and polysome peaks in polysome profile analyses (Figure 5C). To provide mechanistic insight into how the variants impact CDC123 function in human cells, N-terminally His_7_-tagged forms of WT human CDC123 and CDC123-W69C, -N95S, and -V165I were expressed in HEK293T cells and examined for their ability to interact with endogenous eIF2α and eIF2γ.

**Figure 5.**
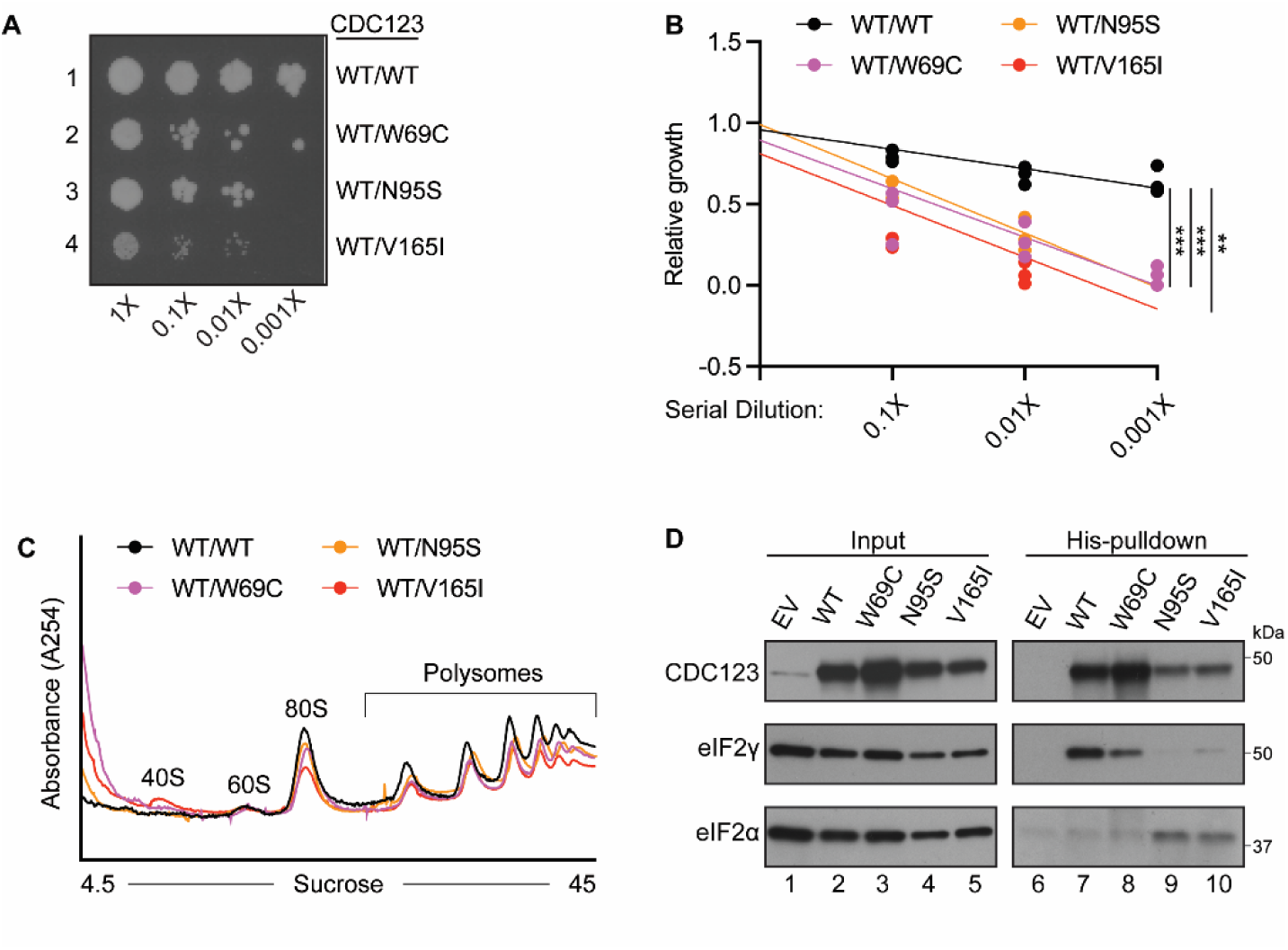
CDC123 variants exhibit dominant-negative impacts on yeast cell growth and disrupt eIF2 subunit interactions in human cells. (A) 10-fold serial dilutions of yeast strains expressing the indicated WT/WT or WT/variant forms of human CDC123 were grown on SD minimal medium at 30°C for 3 days. (B) Linear regression analysis of the relative growth for biological replicate images (n=3) of the spotting assays shown in panel A and Supplemental Figure 8. Statistically significant differences between the slopes of the indicated lines is denoted by an asterisk: **p<0.005, and ***p<0.0005. The slopes and exact p-values for all comparisons shown in panel A are reported in Supplemental Table 1. **(C)** Whole cell extracts (WCEs) from yeast strains expressing the indicated WT/WT or WT/variant forms of CDC123 were subjected to polysome profiling at A254. The 40S and 60S subunits, 80S monosome, and polysome peaks are indicated. **(D)** Whole cell extracts from human HEK293T cells transfected with either an empty vector (EV) or overexpression plasmids for the indicated His_7_-tagged WT or variant forms of CDC123 were either immediately processed for immunoblotting (Input) or incubated with Ni-NTA resin (His-pulldown). Input and pull-down samples were subjected to SDS-PAGE followed by immunoblotting with antibodies against CDC123, eIF2α, or eIF2γ.

Whereas no CDC123 was isolated in an empty vector control, eIF2γ readily co-precipitated with the WT His_7_-tagged CDC123 on Ni^2+^ affinity beads (Figure 5D: compare column 6 vs. column 7). We did not detect eIF2α in our WT CDC123 pull-down, which is consistent with previous reports that eIF2α is rate-limiting for eIF2 complex formation and causes CDC123 dissociation once the eIF2α-eIF2γ heterodimer is formed^33^. Interestingly, substantially less eIF2γ co-precipitated with CDC123-W69C as compared to the wild-type control (Figure 5D). While CDC123-N95S and -V165I may also reduce eIF2γ binding, perhaps more significant was the eIF2α enrichment in the CDC123-N95S and -V165I pull-downs (Figure 5D). Thus, consistent with the results of the CDC123-W69C, -N95S, and -V165I yeast strains, the human CDC123 variants impair CDC123 function by differentially altering the association between CDC123 and the eIF2α and eIF2γ subunits.

## DISCUSSION

In this report, we identify, to our knowledge, the first *CDC123* missense variants to be implicated in human disease. We describe four unrelated families harboring three heterozygous variants in *CDC123* that are classified as likely pathogenic according to guidelines proposed by the American College of Medical Genetics and Genomics and the Association for Molecular Pathology (Figure 1 and Table 1)^35^. While the clinical findings are somewhat variable, potentially owing to differences in the year of clinical assessment and the clinical team involved, shared patient features include developmental abnormalities (*e.g.*, developmental delay, autism, ADHD), hypotonia, and motor dysfunction (Table 1). It should be noted that this manuscript contains all of the available proband and familial information currently available. Paternal exome sequencing from families 1, 3, and 4 (Figure 1A, 1C, and 1D) was not completed, so the inheritance pattern cannot be definitively determined for the associated probands. The fathers in these families were unaffected by a neurodevelopmental phenotype, but paternal sequencing was not conducted nor possible to obtain in these cases. By comparison, maternal and paternal exome sequencing revealed that proband 2 inherited the *CDC123* variant from their father (Figure 1B), whose medical history incudes ADHD, anxiety, and depression which, while consistent with some of the proband’s clinical history, represent a milder set of phenotypes (Table 1). Potential reasons for these phenotypic differences could be that the father displays incomplete penetrance due to unique genetic suppressors or variable expressivity of CDC123-W69C driven by genetic mosaicism. Additional gene expression data and/or familial sequencing will be necessary to address these possibilities in future studies.

Computational assessments of *CDC123* variant frequency in the human population, CDC123 amino acid conservation between species, and CDC123 structural modeling predict that the CDC123-W69C, -N95S, and V165I variants are pathogenic (Figure 2, Figure 3, and Supplemental Figure 5). Immunoblot analysis of protein lysates from CDC123 variant yeast strains demonstrated no difference in CDC123 protein levels, suggesting that these variants do not cause haploinsufficiency (Figure 3I).

Instead, through functional validation we find that the variants are damaging to CDC123 activity. CDC123-W69C, -N95S, and -V165I all confer a slow growth phenotype in yeast that is differentially suppressed by overexpression of eIF2α and eIF2γ (Figure 4A-4C), the two eIF2 subunits that rely on CDC123 activity to promote eIF2α-eIF2γ complex formation^9^. Intriguingly, overexpression of eIF2γ specifically increases cell growth for CDC123-W69C, which maps to a CDC123-eIF2γ interfaces in our structural models (Figure 3A-3F, and Figure 4A). By comparison, only overexpression of eIF2α rescues the slow growth phenotype of CDC123-N95S, while CDC123-V165I is rescued by overexpression of both eIF2α and eIF2γ (Figure 4B-4C). These results are also supported by our AlphaFold multimer model that maps CDC123-N95S to a CDC123-eIF2α interface. CDC123-V165I, however, is centrally located in the protein and could be causing larger CDC123 structural rearrangements that impact both eIF2α and eIF2γ binding (Figure 3A).

Importantly, the probands described here are heterozygous for the reported *CDC123* variants. We, therefore, also developed and assessed the cell growth and protein synthesis levels of humanized yeast strains carrying two CDC123 plasmids. Consistent with a dominant-negative phenotype, strains expressing a WT and a variant CDC123 (-W69C, -N95S, or -V165I) grew markedly slower than the WT/WT CDC123 control (Figure 5A, 5B and Supplemental Figure 8). Polysome profiling analyses further revealed a decrease in the level of actively translating ribosomes in the WT/W69C, WT/N95S, and WT/V165I strains, potentially underlying the observed defect in their cell growth (Figure 5A, 5B, and 5C). From a mechanistic standpoint, previous studies have reported that CDC123 is capable of homodimerization but the precise role of CDC123 dimer formation in the context of eIF2 complex assembly and protein synthesis regulation remains an open question^36^. Finally, CDC123 pull-down experiments in human cells revealed a reduction in the association between eIF2γ and CDC123-W69C, while eIF2α was enriched in the CDC123-N95S and -V165I pull-downs as compared to the WT control (Figure 5D). This result is particularly intriguing as CDC123 is thought to interact with eIF2γ first in the stepwise assembly of the eIF2 complex, and dissociate once the eIF2α-eIF2γ heterodimer is formed^33^. The sustained association of CDC123-N95S and -V165I with eIF2α may suggest that these variants impair eIF2 complex formation by altering the order of CDC123-eIF2 subunit association, whereas CDC123-W69C that maps to an CDC123-eIF2γ interface appears to specifically alter the association of CDC123 with the eIF2γ subunit alone.

A significant impetus for this study was the phenotypic overlap, including developmental and motor delay, between the patients harboring *CDC123* variants and those with MEHMO syndrome, which is caused by eIF2γ variants that reduce levels of active eIF2. Our findings establish a new link between *CDC123* missense variants and eIF2 dysregulation, and further emphasize that CDC123-mediated eIF2 heterotrimer formation is essential for human development. Importantly, this study highlights another example of a growing list of rare, genetic, eIF2-related, neurodevelopmental disorders^7,24,37–42^. It will be crucial to address in future studies why variants in CDC123 and eIF2γ, two proteins that are required for protein synthesis in all cell types, have such significant impacts on nervous system development and function.

## MATERIALS AND METHODS

### Exome sequencing

Exome sequencing was performed on probands and parents (when available) using DNA extracted from blood or buccal swab. Exome library preparation, sequencing, bioinformatics, and data analysis were performed as described previously^43^. Briefly, samples were prepared using either the SureSelect Target Enrichment System (Agilent Technologies, Santa Clara, CA), the Roche NimbleGen EZ Exome System (Madison, WI), or the IDT xGen Exome Research Panel V1.0 (Integrated DNA Technologies, Coralville, Iowa) and sequenced using paired-end, 100 or 150-cycle chemistry on Illumina HiSeq or NextSeq systems (Illumina, San Diego, CA). Stepwise filtering included the removal of common single nucleotide polymorphisms (SNPs), intergenic and 3’/5’ UTR variants, non-splice-related intronic variants, and synonymous variants. Alterations internally classified as pathogenic or likely pathogenic variants, predicted null alterations (nonsense, frameshift, canonical ±1 or 2 splice sites, and initiation codon alterations), and alterations with an HGMD ID were protected from filtering.

### AlphaFold structural modeling

AlphaFold2 (https://cosmic2.sdsc.edu:8443/gateway) was used to generate a CDC123-eIF2α-eIF2γ multimeric structural prediction^31^. The average predicted local distance difference test (pLDDT) scores for structure was approximately 77. The AlphaFold2 structural prediction was visualized using the PyMOL Molecular Graphics System (Version 3.0 Schrödinger).

### Yeast strains and plasmids

Yeast strain YY6 (*Matα ura3-52 trp1-63 leu2-3 leu2-112 GAL2+ CDC123Δ::KanMx p*[Sc-*CDC123, URA3*]) was used for analysis of CDC123 variants. *Sc. and Hs. CDC123* were cloned into *LEU2* expression vectors, and variant alleles of *CDC123* were generated by site-directed mutagenesis of *Hs. CDC123*. All plasmids were sequence verified (Plasmidsaurus), and introduced into YY6 as the sole source of CDC123 by plasmid shuffling.

Plasmids used in the eIF2 subunit overexpression assays were generated using a NEBuilder HiFi DNA Assembly kit. Briefly, *TRP1* was homologously recombined with the DNA fragments generated from empty vector, eIF2α, or eIF2γ-containing *Yep24* plasmids to generate either an empty vector plasmid, or eIF2α and eIF2γ overexpression plasmids with a *TRP1* selection marker.

For the strains expressing CDC123 from two plasmids, a plasmid encoding human WT CDC123 with a *URA3* selectable marker was transformed into strains harboring human WT CDC123, -W69C, -N95S, or -V165I plasmids with a *LEU2* marker. All plasmids used in this study were verified by sequencing.

### Growth analyses

Derivatives of yeast strain YY6 expressing *Sc* WT CDC123, *Hs* WT CDC123, or variant forms of human CDC123, either with an empty vector or high copy plasmids to overexpress yeast eIF2α (*SUI2)* or eIF2γ (*GCD11*) were colony purified on synthetic defined (SD) minimal medium supplemented with amino acids (+AA). Similarly, humanized yeast strains derived from YY6 and expressing *Hs* WT CDC123, CDC123-W69C, -N95S, or -V165I from *LEU2* plasmids were co-transformed with a *URA3* plasmid expressing Hs WT CDC123 and colony purified on SD minimal medium +AA. A single colony from each strain was used to inoculate 225 μL of UltraPure Water and subsequently vortexed. Absorbance measurements were conducted and samples were normalized and diluted to generate 10-fold serial dilutions (OD_600_ = 1.0, 0.1, 0.01, 0.001). 3 μL volumes of the serial dilutions were spotted onto SD medium containing the required amino acid supplements, and the plates were incubated at 30°C for 3 days prior to imaging.

### Polysome profiling analyses

Humanized yeast strains derived from YY6 and expressing *Hs* WT CDC123, CDC123-W69C, -N95S, or -V165I from *LEU2* plasmids were co-transformed with a *URA3* plasmid expressing Hs WT CDC123 and colony purified on SD minimal medium +AA. A single colony from each strain was used to inoculate 40 mL of SD liquid medium +AA that was grown with shaking at 30°C for 24 hours. 20 μL of the starter culture was used to inoculate 50 mL of fresh SD liquid medium + AA and the cells were incubated at 30°C with shaking for 16 hours until the cultures reached mid-exponential growth phase (OD_600_ ∼ 1). Cells were transferred to pre-chilled 50 mL conical tubes on ice containing 500 μL of 10 mg/mL cycloheximide (CHX) and centrifuged at 5,000 RPM for 5 minutes at 4°C. The supernatant was aspirated and cells were rinsed with 25 mL chilled 1X lysis buffer (20 mM Tris-HCl pH 7.4, 50 mL KCl, 10 mM MgCl_2_, 1 mM DTT, and 0.1 mg/mL CHX) before being promptly centrifuged at 5,000 RPM for 5 minutes at 4°C. The supernatant was aspirated and cells were resuspended in 800 μL of 1X lysis buffer and transferred to a microcentrifuge tube on ice. The cells were pelleted by centrifugation at 5,000 RPM for 5 minutes at 4°C and the supernatant was discarded. Cells were resuspended in 300 μL of 1X lysis buffer and transferred to pre-chilled flat-bottomed microcentrifuge tubes on ice containing 300 μL of acid-washed beads. In the cold room, the cells were vortexed for 20 seconds and cooled in ice water for 40 seconds for a total of 6 times. The lysate was clarified by centrifugation at 10,000 RPM for 5 minutes at 4°C, transferred to a new pre-chilled microcentrifuge tube, and clarified by centrifugation at 10,000 RPM for 5 minutes at 4°C a second time. The supernatant was recovered and transferred to a new pre-chilled microcentrifuge tube on ice. The samples were measured at OD_260_ and lysate concentrations were equilibrated in a total volume of 350 μL. 300 μL of each equilibrated cell lysate was layered on a 4.5 to 45% sucrose gradient containing 20 mM Tris-HCl pH 7.4, 50 mL KCl, 10 mM MgCl_2_, and 0.1 mg/mL CHX. Sucrose gradient preparation, ultracentrifugation, and polysome profile collection were conducted as described in ^44^.

### Cell culture and transfection

HEK293T cells were maintained in Dulbecco Modified Eagle Medium (DMEM) containing 4.5 g/L D-glucose, L-glutamine and 110 mg/L sodium pyruvate (Gibco) that was supplemented with 10% Fetal Bovine Serum and 1% penicillin/streptomycin. One day prior to transfection, cells were seeded at 0.21 × 10^6^ in 6-well plates and grown at 37°C in 5% CO_2_. For CDC123 pull-down analyses (described below), an empty vector or overexpression plasmids encoding N-terminal His_7_-tagged WT or variant CDC123 were transiently transfected into HEK293T cells following the Lipofectamine 3000 (Invitrogen) manufacturer’s protocol. Culture media was exchanged for fresh DMEM medium 6 hours post transfection. Cells were allowed to grow for an additional 30 hours before cell lysis (described below).

### Immunoblot analysis

Yeast strains expressing WT or variant forms of CDC123 were grown to OD_600_∼1 in 200 ml SD medium containing required amino acid supplements at 30°C. Cells were harvested at 3,000 rpm for 5 minutes, washed with 10 ml Breaking Buffer B (20 mM Tris–HCl (pH 7.5), 500 mM KCl, 0.1 mM MgCl_2_, 10 mM imidazole, 5 mM β-mercaptoethanol, 10% (v/v) glycerol, 1× cOmplete protease inhibitor, and 1 mM 4-(2-aminoethyl) benzenesulfonyl fluoride hydrochloride (AEBSF), and resuspended in 2 ml Breaking Buffer B. Cells were lysed by adding 50% (w/v) glass beads followed by vigorous mixing on a vortex for 5 min at 4°C. Lysates were cleared by centrifugation at 14,000 × *g* for 10 min at 4°C, quantified, mixed with sodium dodecyl sulphate (SDS)-sample buffer with dithiothreitol (DTT) and incubated at 70°C for 10 min. Samples were subjected to SDS-polyacrylamide gel electrophoresis (SDS-PAGE) followed by immunoblotting with rabbit polyclonal antiserum against human CDC123 (Genscript) and yeast eIF2β^45^.

### CDC123 pull-down and immunoblot analysis

For cell lysis of transfected HEK293T cells, DMEM was aspirated and the cells were washed with 2 mL of chilled 1X PBS per well. 2.5 μL lysis buffer (50 mM NaH_2_PO_4_, 300 mM NaCl, 10 mM imidazole, 0.05% Tween 20, pH 8.0) was added to each well and cells were subsequently collected with a cell scraper. The lysate was then transferred to a microcentrifuge tube, incubated on ice for 5 minutes, and cell debris was pelleted by centrifugation at 13,000 RPM for 10 minutes at 4°C. The supernatant was collected into a new microcentrifuge tube on ice, and the protein concentration of each lysate was determined using diluted Bio-Rad Protein Assay Dye Reagent Concentrate per the manufacturer’s protocol. For protein input samples, one quarter of the lysate was normalized, mixed with SDS-sample buffer, and incubated at 70°C for 10 minutes.

For CDC123 pull-down assays, extracts were normalized for protein concentration by diluting the more concentrated extracts with lysis buffer to match the protein concentration of the least concentrated extract in a total volume of 800 μL. Lysates were then mixed with 0.1 mL Ni-NTA resin (Qiagen), and incubated on a Nutator mixer at 4°C for 1 hour. Following nutation, the Ni-NTA beads were washed three times with 10 mL of wash buffer (50 mM NaH_2_PO_4_, 300 mM NaCl, 20 mM imidazole, 0.05% Tween 20, pH 8.0). Beads were resuspended in 1 mL wash buffer and transferred to microcentrifuge tubes that were subsequently centrifuged at 300 × g for 5 minutes. The supernatant was removed and beads were resuspended in 45 μL of elution buffer (50 mM NaH_2_PO_4_, 300 mM NaCl, 250 mM imidazole, 0.05% Tween 20, pH 8.0) and incubated for 1 minute at room temperature. Beads were then centrifuged at 300 × g for 5 minutes and equal volumes of supernatant were mixed with SDS-sample buffer and heated at 70°C for 10 minutes. Proteins bound to the resin were analyzed by 10% SDS-PAGE and immunoblotting with antibodies against CDC123 (GenScript), eIF2α (Abcam), or eIF2γ (GenScript). The CDC123 and eIF2γ primary antibodies were generated against recombinant CDC123 and eIF2γ proteins and tested for reactivity against their respective antigen by Genscript.

### Quantification and statistical analysis

For the yeast cell spotting assay quantifications, biological replicate (n=3) images were inverted in Photoshop and the integrated density of individual spots was determined using ImageJ. Data from individual spots were normalized to measurement from the first spot for each yeast strain’s serial dilution series. GraphPad Prism (Version 11) was used to conduct simple linear regression with a multiple comparisons test. Significance between slopes from the lines of best fit is indicated by an asterisk: *p<0.05, **p<0.005, ***p<0.0005, ns = not significant.

## DATA AVAILABILITY

The data underlying this article are available in the article and in its online supplementary material. The DNA sequence variants are available on ClinVar, at https://www.ncbi.nlm.nih.gov/clinvar/.

## ACKNOWLEDGMENTS

The authors are indebted to the patients and families for their cooperation.

## CReDit STATEMENT WITH AUTHOR CONTRIBUTIONS

A.L.E.: Writing – original draft, Visualization, Validation, Methodology, Investigation, Formal analysis, Data curation, Conceptualization.

A.M.: Writing – review and editing,Validation, Investigation, Data curation.

A.R-M.: Writing – review and editing, Validation, Investigation.

B.G.: Writing – review and editing, Data curation.

G.E.V.: Writing – review and editing, Supervision, Data curation.

C.D.: Writing – review and editing, Investigation, Data curation.

A.E.G.: Writing – review and editing, Investigation, Data curation.

D.B.: Writing – review and editing, Supervision, Investigation, Data curation.

S.K.Y.-B.: Writing – original draft, Supervision, Resources, Project administration, Methodology, Funding acquisition, Formal analysis, Data curation, Conceptualization.

## ETHICS DECLARATION

Per the determination of the Western Institutional Review Board (WIRB), the research reported in this manuscript does not constitute human subjects research and is exempt from formal IRB review. Informed consent is not required as part of the IRB waiver. This determination was made by the WIRB based on federal regulation 45 CFR 46 and associated guidance. All data reported in the manuscript are de-identified.

## REFERENCES

1. Young, S.K. & Wek, R.C. Upstream Open Reading Frames Differentially Regulate Gene-specific Translation in the Integrated Stress Response. J Biol Chem 291, 16927–35 (2016).

2. Wek, R.C. Role of eIF2alpha Kinases in Translational Control and Adaptation to Cellular Stress. Cold Spring Harb Perspect Biol 10, a032870 (2018).

3. Wek, R.C., Anthony, T.G. & Staschke, K.A. Surviving and Adapting to Stress: Translational Control and the Integrated Stress Response. Antioxid Redox Signal (2023).

4. Ron, D. Translational control in the endoplasmic reticulum stress response. J Clin Invest 110, 1383–8 (2002).

5. Costa-Mattioli, M. & Walter, P. The integrated stress response: From mechanism to disease. Science 368(2020).

6. Tahmasebi, S., Khoutorsky, A., Mathews, M.B. & Sonenberg, N. Translation deregulation in human disease. Nat Rev Mol Cell Biol 19, 791–807 (2018).

7. Marintchev, A. & Ito, T. eIF2B and the Integrated Stress Response: A Structural and Mechanistic View. Biochemistry 59, 1299–1308 (2020).

8. Hinnebusch, A.G. Molecular mechanism of scanning and start codon selection in eukaryotes. Microbiol Mol Biol Rev 75, 434–67 (2011).

9. Perzlmaier, A.F., Richter, F. & Seufert, W. Translation initiation requires cell division cycle 123 (Cdc123) to facilitate biogenesis of the eukaryotic initiation factor 2 (eIF2). J Biol Chem 288, 21537–46 (2013).

10. Erb, A.L. & Young-Baird, S.K. CDC123 is an ATPase that modulates mRNA translation and the integrated stress response by regulating eIF2 complex assembly. J Biol Chem 302, 111116 (2025).

11. Bieganowski, P., Shilinski, K., Tsichlis, P.N. & Brenner, C. Cdc123 and checkpoint forkhead associated with RING proteins control the cell cycle by controlling eIF2γ abundance. J Biol Chem 279, 44656–66 (2004).

12. Sonenberg, N., Hershey, J.W.B. & Mathews, M.B. Translational control of gene expression, (Cold Spring Harbor Laboratory Press, Cold Spring Harbor, N.Y, 2000).

13. Llacer, J.L. et al. Conformational differences between open and closed states of the eukaryotic translation initiation complex. Mol Cell 59, 399–412 (2015).

14. Marintchev, A. Exploring the interaction dynamics of eukaryotic translation initiation factor 2. Biochem Soc Trans 53, 593–602 (2025).

15. Young-Baird, S.K. et al. Suppression of MEHMO Syndrome Mutation in eIF2 by Small Molecule ISRIB. Mol Cell 77, 875–886 e7 (2020).

16. Shiber, A. et al. Cotranslational assembly of protein complexes in eukaryotes revealed by ribosome profiling. Nature 561, 268–272 (2018).

17. DeLozier-Blanchet, C.D., Haenggeli, C.A. & Bottani, A. MEHMO, a novel syndrome: assignment of disease locus to Xp21.1-p22.13. Mental retardation, epileptic seizures, hypogonadism and genitalism, microcephaly, obesity. Eur J Hum Genet 7, 621–2 (1999).

18. Steinmuller, R., Steinberger, D. & Muller, U. MEHMO (mental retardation, epileptic seizures, hypogonadism and -genitalism, microcephaly, obesity), a novel syndrome: assignment of disease locus to xp21.1-p22.13. Eur J Hum Genet 6, 201–6 (1998).

19. Borck, G. et al. eIF2γ mutation that disrupts eIF2 complex integrity links intellectual disability to impaired translation initiation. Mol Cell 48, 641–6 (2012).

20. Skopkova, M., et al. *EIF2S3* Mutations Associated with Severe X-Linked Intellectual Disability Syndrome MEHMO. Hum Mutat 38, 409–425 (2017).

21. Gregory, L.C. et al. Impaired EIF2S3 function associated with a novel phenotype of X-linked hypopituitarism with glucose dysregulation. EBioMedicine 42, 470–480 (2019).

22. Young-Baird, S.K., Shin, B.S. & Dever, T.E. MEHMO syndrome mutation EIF2S3-I259M impairs initiator Met-tRNAiMet binding to eukaryotic translation initiation factor eIF2. Nucleic Acids Res 47, 855–867 (2019).

23. Kotzaeridou, U. et al. Novel pathogenic EIF2S3 missense variants causing clinically variable MEHMO syndrome with impaired eIF2gamma translational function, and literature review. Clin Genet 98, 507–514 (2020).

24. Dang Do, A.N., Navid, F. & Young-Baird, S.K. MEHMO syndrome: Review and proposed classification as an eIF2-related neuroendocrinopathy. Mol Genet Metab 146, 109287 (2025).

25. Moortgat, S. et al. Two novel *EIF2S3* mutations associated with syndromic intellectual disability with severe microcephaly, growth retardation, and epilepsy. Am J Med Genet A 170, 2927–2933 (2016).

26. Moortgat, S. et al. Broadening the phenotypic spectrum and physiological insights related to EIF2S3 variants. Hum Mutat 42, 827–834 (2021).

27. Mori, M. et al. Ketogenic diet for refractory epilepsy with MEHMO syndrome: Caution for acute necrotizing pancreatitis. Brain Dev 43, 724–728 (2021).

28. Ivanova, N., Serzhanova, V., Demina, N., Guseva, D. & Skoblov, M. mRNA analysis revealed a novel pathogenic EIF2S3 variant causing MEHMO syndrome. Eur J Med Genet 65, 104421 (2022).

29. Frederic, M.Y. et al. UMD-predictor, a new prediction tool for nucleotide substitution pathogenicity -- application to four genes: FBN1, FBN2, TGFBR1, and TGFBR2. Hum Mutat 30, 952–9 (2009).

30. Suybeng, V., Koeppel, F., Harle, A. & Rouleau, E. Comparison of Pathogenicity Prediction Tools on Somatic Variants. J Mol Diagn 22, 1383–1392 (2020).

31. Jumper, J. et al. Highly accurate protein structure prediction with AlphaFold. Nature 596, 583–589 (2021).

32. Panvert, M. et al. Cdc123, a Cell Cycle Regulator Needed for eIF2 Assembly, Is an ATP-Grasp Protein with Unique Features. Structure 23, 1596–1608 (2015).

33. Vanselow, S., Neumann-Arnold, L., Wojciech-Moock, F. & Seufert, W. Stepwise assembly of the eukaryotic translation initiation factor 2 complex. J Biol Chem 298, 101583 (2022).

34. Cardenal Peralta, C., et al. Binding of human Cdc123 to eIF2gamma. J Struct Biol 215, 108006 (2023).

35. Richards, S. et al. Standards and guidelines for the interpretation of sequence variants: a joint consensus recommendation of the American College of Medical Genetics and Genomics and the Association for Molecular Pathology. Genet Med 17, 405–24 (2015).

36. Bertolini, M. et al. Interactions between nascent proteins translated by adjacent ribosomes drive homomer assembly. Science 371, 57–64 (2021).

37. Mao, D. et al. De novo EIF2AK1 and EIF2AK2 Variants Are Associated with Developmental Delay, Leukoencephalopathy, and Neurologic Decompensation. Am J Hum Genet 106, 570–583 (2020).

38. Kuipers, D.J.S. et al. EIF2AK2 Missense Variants Associated with Early Onset Generalized Dystonia. Ann Neurol 89, 485–497 (2021).

39. Macintosh, J., Thiffault, I., Pastinen, T., Sztriha, L. & Bernard, G. A Recurrent De Novo Variant in EIF2AK2 Causes a Hypomyelinating Leukodystrophy. Child Neurol Open 10, 2329048X231176673 (2023).

40. Delepine, M., et al. *EIF2AK3*, encoding translation initiation factor 2-α kinase 3, is mutated in patients with Wolcott-Rallison syndrome. Nature Genetics 25, 406–409 (2000).

41. Abdulkarim, B. et al. A Missense Mutation in PPP1R15B Causes a Syndrome Including Diabetes, Short Stature, and Microcephaly. Diabetes 64, 3951–62 (2015).

42. Kernohan, K.D. et al. Homozygous mutation in the eukaryotic translation initiation factor 2alpha phosphatase gene, PPP1R15B, is associated with severe microcephaly, short stature and intellectual disability. Hum Mol Genet 24, 6293–300 (2015).

43. Farwell, K.D. et al. Enhanced utility of family-centered diagnostic exome sequencing with inheritance model-based analysis: results from 500 unselected families with undiagnosed genetic conditions. Genet Med 17, 578–86 (2015).

44. Rodriguez-Martinez, A. & Young-Baird, S.K. Polysome profiling is an extensible tool for the analysis of bulk protein synthesis, ribosome biogenesis, and the specific steps in translation. Mol Biol Cell 36, mr2 (2025).

45. Dever, T.E., Yang, W., Åström, S., Byström, A.S. & Hinnebusch, A.G. Modulation of tRNA_i_^Met^, eIF-2 and eIF-2B expression shows that *GCN4* translation is inversely coupled to the level of eIF-2.GTP.Met-tRNA_i_^Met^ ternary complexes. Mol Cell Biol 15, 6351–6363 (1995).

