## Supplementary Material for "CDC123 variants associated with developmental abnormalities alter CDC123-eIF2 subunit interactions and impair protein synthesis"

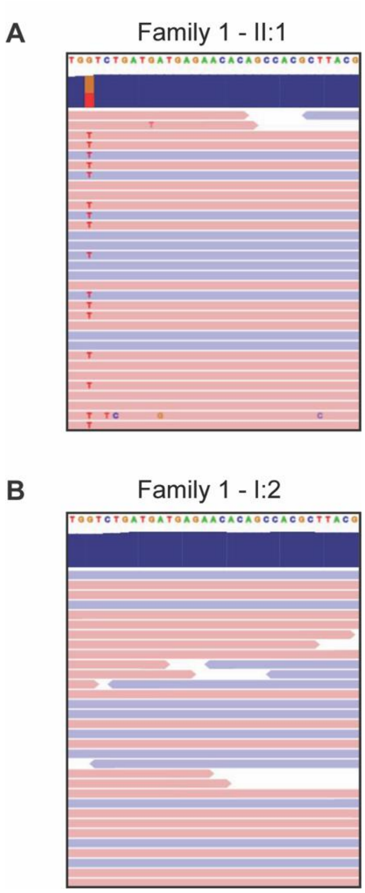


**Supplemental Figure 1. Identification of the *CDC123* c.207G>T (p.W69C) variant for Family 1.** Expanded IGV displays of exonic sequence data for **(A)** proband 1 and **(B)** their mother. Blue and red bars indicate individual sequencing reads from the positive (blue) and negative (red) DNA strands. *CDC123* variant sequencing reads display a red “T” in the blue and red bars. The WT *CDC123* sequence is shown at the top of the IGV images.

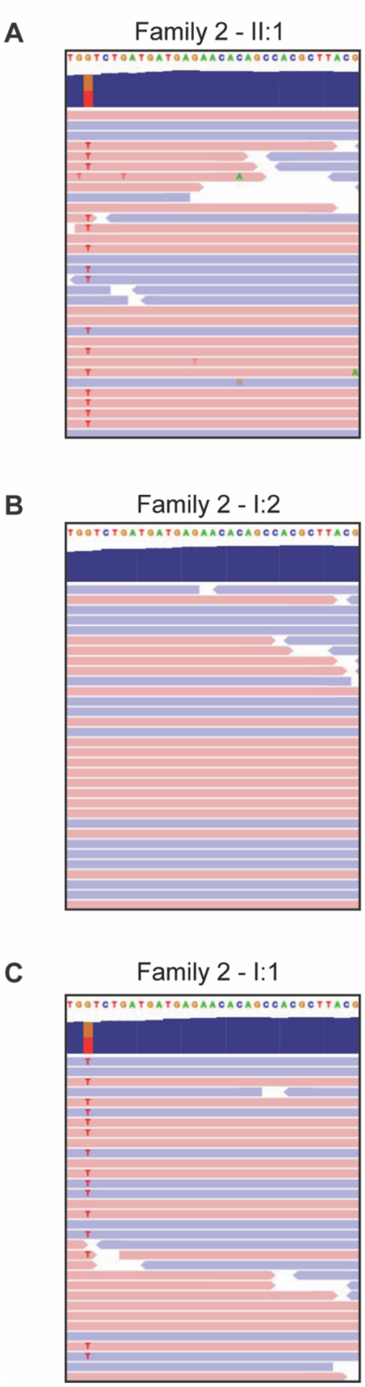


**Supplemental Figure 2. Identification of the *CDC123* c.207G>T (p.W69C) variant for Family 2.** Expanded IGV displays of exonic sequence data for **(A)** proband 2, **(B)** their mother, and **(C)** their father. Blue and red bars indicate individual sequencing reads from the positive (blue) and negative (red) DNA strands. *CDC123* variant sequencing reads display a red “T” in the blue and red bars. The WT *CDC123* sequence is shown at the top of the IGV images.

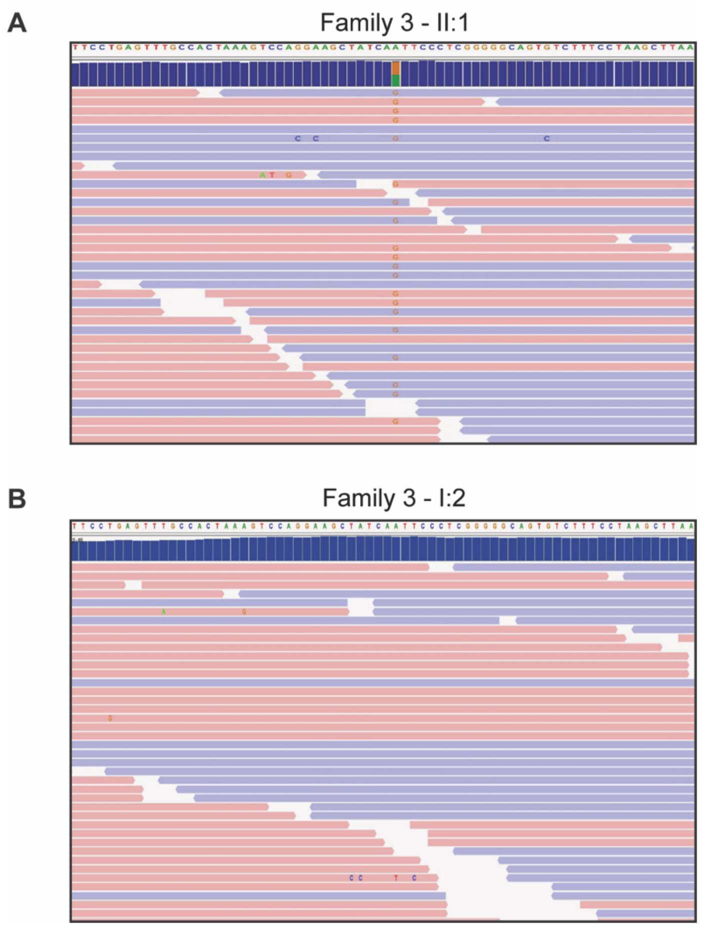


**Supplemental Figure 3. Identification of the *CDC123* c.284A>G (p.N95S) variant for Family 3.** Expanded IGV views of exonic sequence data for **(A)** proband 3 and **(B)** their mother. Blue and red bars indicate individual sequencing reads from the positive (blue) and negative (red) DNA strands. *CDC123* variant sequencing reads display an orange "G" in the middle of the blue and red bars. The WT *CDC123* sequence is shown at the top of the IGV images.

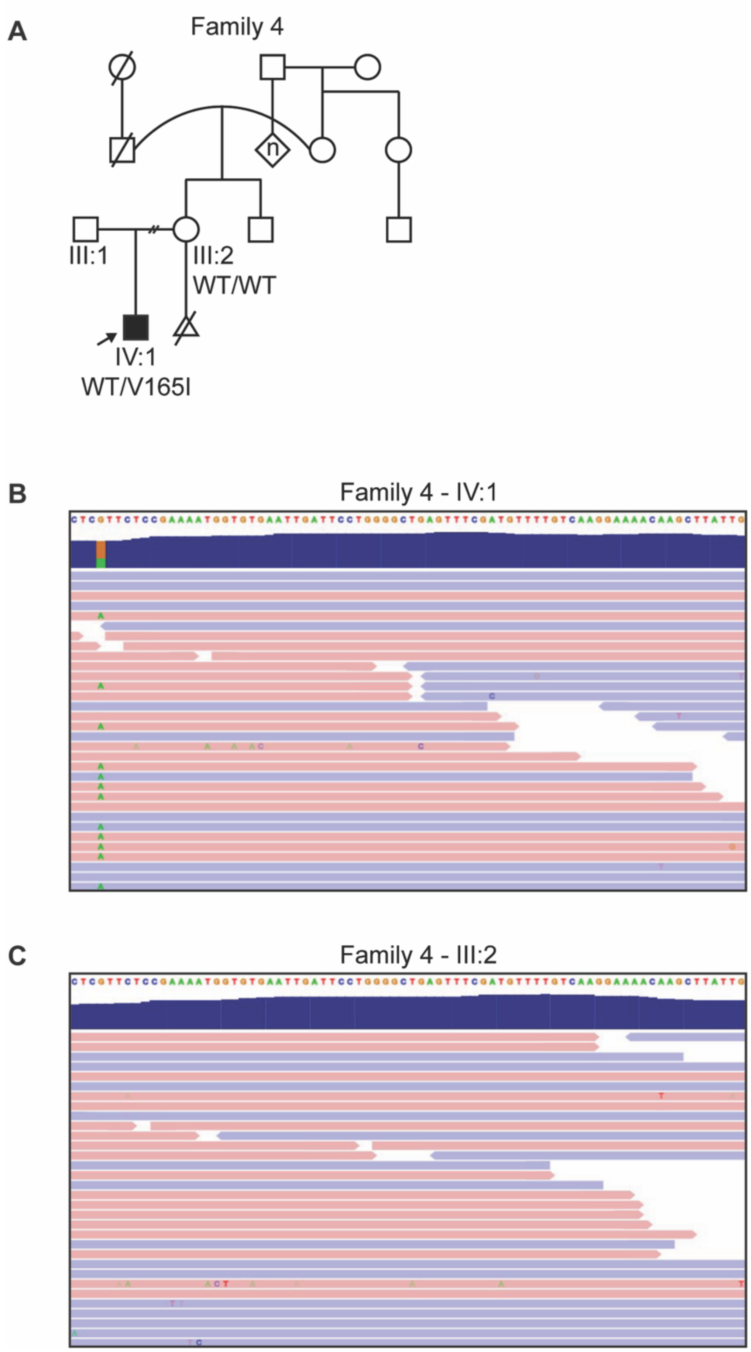


**Supplemental Figure 4. Identification of the *CDC123* c.493G>A (p.V165I) variant for Family 4. (A)** Expanded pedigree analysis for Family 4. Individuals affected by developmental abnormalities are shown as filled symbols in the familial pedigree. Additional symbols denote the following: open square = unaffected male, open circle =

unaffected female, diamond = unknown biological sex, n = unknown number of children, and triangle = miscarriage. The proband is indicated by an arrow. **(B)** Expanded IGV displays of exonic sequence data for proband 4 and **(C)** their mother. Blue and red bars indicate individual sequencing reads from the positive (blue) and negative (red) DNA strands. *CDC123* variant sequencing reads display a green “A” in the blue and red bars. The WT *CDC123* sequence is shown at the top of the IGV images.

```
H. sapiens      -------------MKKEHVLHCQFSAWYPFFRGVTIKSVILP-LPQNVKDYLLDDGTLVVSGRDDPPTHSQPDSDDE
B. taurus       -------------MKKEHVLHCQFSAWYPLFRSLTIKSVILP-LPQNVKDYLLDDGTLVVSGREDPPAHSQPDSDDE
S. scrofa       -------------MKKEHVLHCQFSAWYPLFRSLTIKSVILP-LPQNVKDYLLDDGTLVVSGREDPPARSQPDSDDE
R. norvegicus   -------------MKKEHVSHCQFSAWYPLFRSLTIKSVILP-LPQNVKDYLLDDGTLVVSGREDPPTCSQPDSGDE
M. musculus     -------------MKKEHVSHCQFSAWYPLFRSLTIKSVILP-LPQNVKDYLLDDGTLVVSGREDPPTCSQSDSGNE
S. cerevisiae   MSSQEYTTFIDIPVTRAQVEHCSYSFWSSLYPKYVPKSIVLKSLPKKFIQYLEQDGIKLPQEENSRSVYTEEIIRNE
                             :.: :* **.:* * .::   . **::*  **::. :** :**  : . .:. .. ::    :*

H. sapiens      -AEEIQWSDDENTATLTAP------EFPEFATKVQEAINSLGGSVFPKLNWSAPRDAYWIAMNSSLKCKTLSDIFLL
B. taurus       -AEEIQWSDDENTATLTAP------EFPEFTTKVQEAINSLGGSVFPKLNWSAPRDAYWIAMNSSLKCKTLSDIFLL
S. scrofa       -AEEIQWSDDENTATLTAP------EFPEFTTKVQEAINSLGGSVFPKLNWSAPRDAYWIAMNSSLKCKTLSDIFLL
R. norvegicus   -AEETQWSDDESTATLTAP------EFPEFNTQVQEAINSLGGSVFPKLNWSAPRDAYWIAMNSSLKCKSLSDIFLL
M. musculus     -AEETQWSDDESTATLTAP------EFPEFNTQVQEAINSLGGSVFPKLNWSAPRDAYWIAMNSSLKCKTLSDIFLL
S. cerevisiae   DNDYSDWEDDEDTATEFVQEVEPLIDFPELHQKLKDALNELG-AVAPKLNWSAPRDATWILPNNTMKCNEVNELYLL
                  :  :*.***.***  .       :***:  ::::*:*.** :* *********** **  *.::**: :.:::**

H. sapiens      FKSSDFITRDFTQPFIHCTDDSPDPCIEYELVLRKWCELIPGAEFRCFVKENKLIGISQRDYTQYYDHISKQKEEIR
B. taurus       FKSSDFITRDFTQPFIHCTDDSPDPCMEYELVLRKWCELIPGAEFRCFVKENKLIGISQRDYTQYYDHISKQKEEIC
S. scrofa       FKSSDFITRDFTQPFIHCTDDSPDPCMEYELVLRKWCELIPGAEFRCFVKENKLIGISQRDYTQYYDHISKQKEEIC
R. norvegicus   FKSSDFITHDFTQPFIHCNDDSPDPCIEYELVLRKWCELIPGAEFRCFVKENKLIGISQRDYTQYYDHISKQKEEIC
M. musculus     FKSSDFITHDFTQPFIHCTDDSPDPCIEYELVLRKWCELIPGAEFRCFVKENKLIGISQRDYTQYYDHISKQKEEIC
S. cerevisiae   LNASNYIMHDLQRAFKGCVDGDDIKGLKFDLVLRQWCDMNPALEFRVFVKNAHIVGATQRDLN-YYDYLDELSDTFK
                :::*::* :*: :.*  * *..    ::::****:**:: *. *** ***: :::* :*** . ***::.: .: :

H. sapiens      RCIQDFFKKHIQYKFLDEDFVFDIYRD-SRGKVWLIDFNPFGEVTDSLLFTWEELISENNLNG---DFSEVDAQEQD
B. taurus       RCIQDFFKKHIQYKFLDEDFVFDIYRD-SRGKVWLIDFNPFGEVTDSLLFTWDELLSGTNLKG---DFS-EEALEQD
S. scrofa       RCIQDFFKKHIQYKFLDEDFVFDIYRD-SRGKVWLIDFNPFGEVTDALLFTWEELLSGRSVRG---DLSEGEAPEQD
R. norvegicus   RCIQDFFKEHLQYKFLDEDFVFDIYRD-SRGKVWLIDFNPFGEVTDSLLFTWEELTSENNLRG---DVSEADALEQD
M. musculus     RCIQDFFKEHLQYKFLDEDFVFDIYRD-SRGKVWLIDFNPFGEVTDSLLFTWEELTSENNLRG---EVTEGDAQEQD
S. cerevisiae   DLIDEIVHDVVLPKFPDKSFVLDVYIPRPFNKIFIVDINPFARKTDSLLFSWNEIAAIAPPKNDVEDYELRLVTRHN
                  *:::.:. :  ** *:.**:*:*   . .*::::*:***.. **:***:*:*: :    ..   :     . .::

H. sapiens      SPAFRCTNSEVTVQPSPYLSYRLPKDFVDLSTGEDAHKLIDFLKLKRNQQEDD-
B. taurus       APAFRCTNSEVTVQPSPYLSYRLPKDFVDLSTGEDAHKLIDFLKLKRNQQEDD-
S. scrofa       APTFRCTNSEVTVQPSPYLSYRLPKDFVDLSTGEDAHKLIDFLKLKRNQQEDD-
R. norvegicus   SPAFRCTNSEVTVQPSPYLSYGLPKDFVDLSTGEDAHKLIDFLKLKRNQQEDD-
M. musculus     SPAFRCTNSEVTVQPSPYLSFGLPKDFVDLSTGEDAHKLIDFLKLKRNEQEDD-
S. cerevisiae   TGRFASKEHSENHVPQDLVEASLNPEAIRELTQKWKELLSQQAKEESSDSENET
                :  * ..: . .  *.  :.  *  : :   * :  . * :  * : .:.*::
```

**Supplemental Figure 5. CDC123-W69, -N95, and -V165 are highly conserved amino acid residues.** Multiple sequence alignment of CDC123 amino acid sequences from *H. sapiens*, *B. taurus*, *S. scrofa, R. norvegicus, M. musculus,* and *S. cerevisiae.* CDC123-W69 (purple), -N95 (orange), and -V165I (red) are located within highly conserved amino acid motifs. Fully conserved, strongly similar, weakly similar, and non-conserved residues are indicated by an asterisk (*), semicolon (:), period (.), and no symbol, respectively.

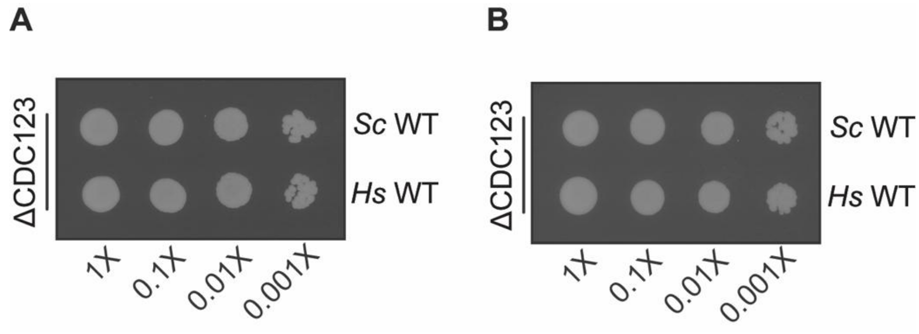


**Supplemental Figure 6. Biological replicate images used for the spotting assay quantification in Figure 3H.**

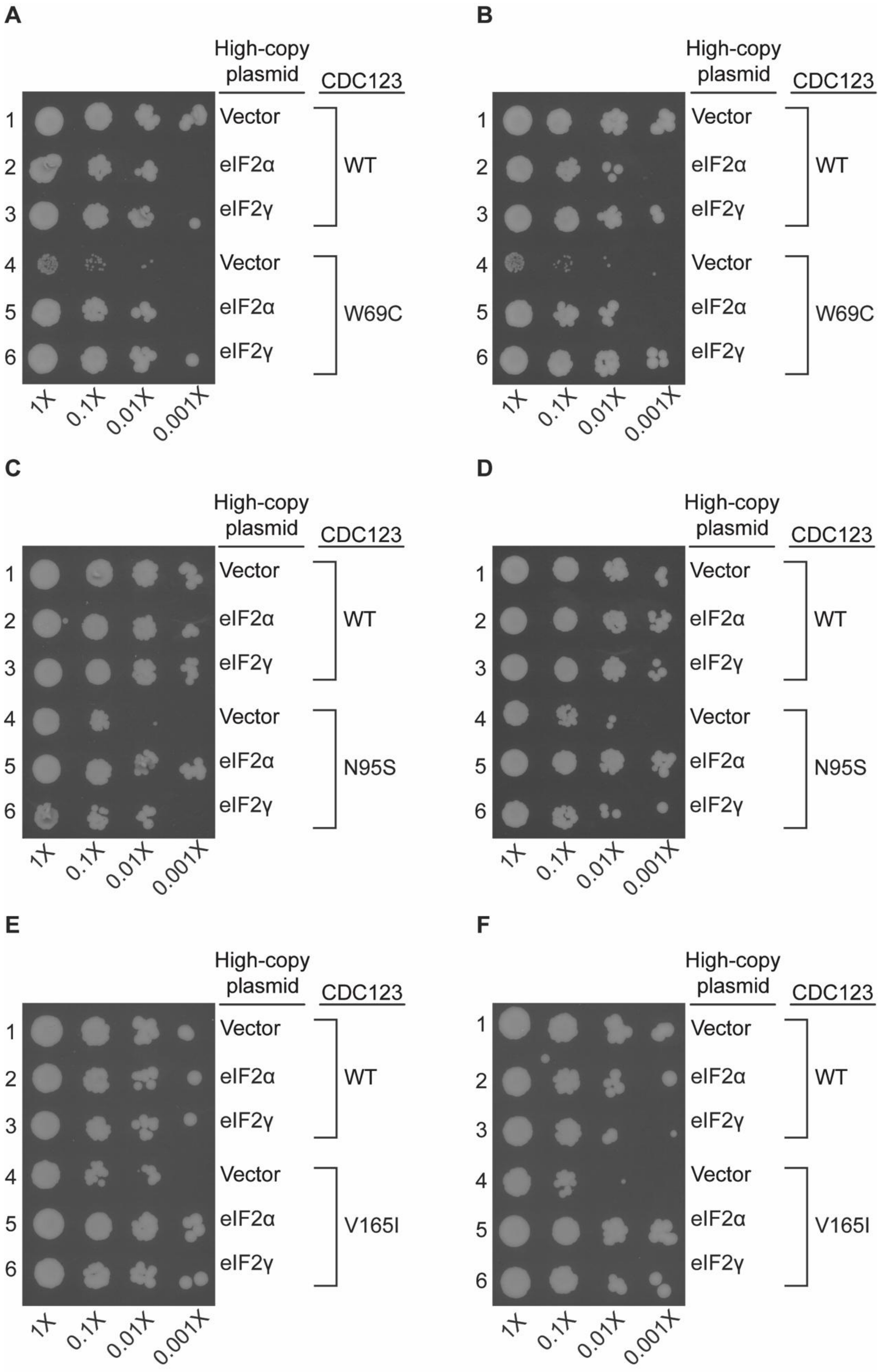

A
High-copy plasmid
CDC123
1
2
3
4
5
6
Vector
eIF2α
eIF2γ
Vector
eIF2α
eIF2γ
WT
W69C
1X
0.1X
0.01X
0.001X
B
High-copy plasmid
CDC123
1
2
3
4
5
6
Vector
eIF2α
eIF2γ
Vector
eIF2α
eIF2γ
WT
W69C
1X
0.1X
0.01X
0.001X
C
High-copy plasmid
CDC123
1
2
3
4
5
6
Vector
eIF2α
eIF2γ
Vector
eIF2α
eIF2γ
WT
N95S
1X
0.1X
0.01X
0.001X
D
High-copy plasmid
CDC123
1
2
3
4
5
6
Vector
eIF2α
eIF2γ
Vector
eIF2α
eIF2γ
WT
N95S
1X
0.1X
0.01X
0.001X
E
High-copy plasmid
CDC123
1
2
3
4
5
6
Vector
eIF2α
eIF2γ
Vector
eIF2α
eIF2γ
WT
V165I
1X
0.1X
0.01X
0.001X
F
High-copy plasmid
CDC123
1
2
3
4
5
6
Vector
eIF2α
eIF2γ
Vector
eIF2α
eIF2γ
WT
V165I
1X
0.1X
0.01X
0.001X

**Supplemental Figure 7. Biological replicate images used for the spotting assay quantification in Figures 4D, 4E, and 4F.**

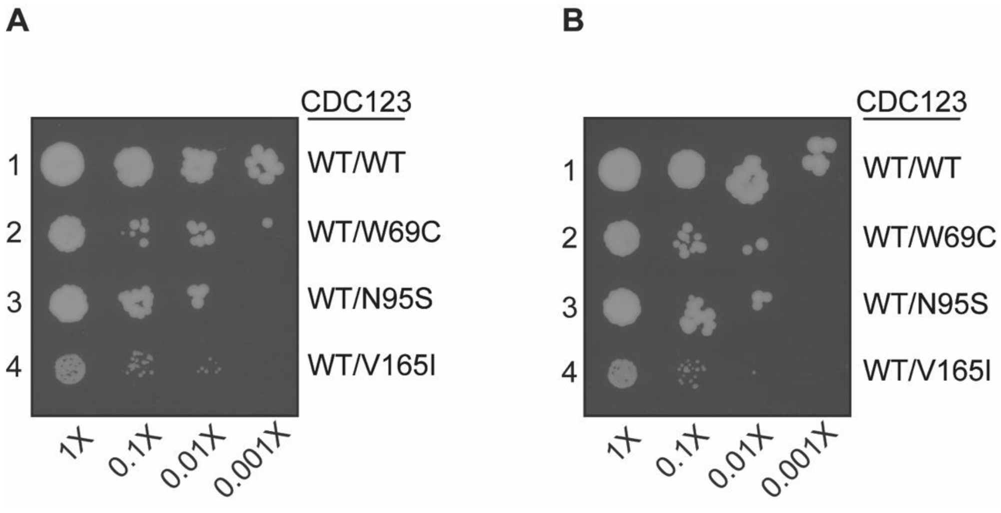


**Supplemental Figure 8. Biological replicate images used for the spotting assay quantification in Figure 5B.**

| Figure reference | Test group | Slope | Comparison | p-value (Are the slopes between two lines different?) |
|---|---|---|---|---|
| 3H | *Sc*-WT CDC123 | -0.0880 | | |
| 3H | *Hs*-WT CDC123 | -0.0924 | *Sc*-WT CDC123 | ns (p=0.7843) |
| 4D | WT Empty Vector | -0.1145 | | |
| 4D | W69C Empty Vector | -0.3208 | WT Empty Vector | *** (p=0.0004) |
| 4D | W69C-eIF2α | -0.2667 | W69C Empty Vector | ns (p=0.3086) |
| 4D | W69C-eIF2γ | -0.1833 | W69C Empty Vector | * (p=0.0278) |
| 4E | WT Empty Vector | -0.1827 | | |
| 4E | N95S Empty Vector | -0.3496 | WT Empty Vector | *** (p=0.0002) |
| 4E | N95S-eIF2α | -0.1250 | N95S Empty Vector | *** (p<0.0001) |
| 4E | N95S-eIF2γ | -0.3188 | N95S Empty Vector | ns (p=0.4817) |
| 4F | WT Empty Vector | -0.2120 | | |
| 4F | V165I Empty Vector | -0.3421 | WT Empty Vector | *** (p=0.0004) |
| 4F | V165I-eIF2α | -0.0933 | V165I Empty Vector | *** (p<0.0001) |
| 4F | V165I-eIF2γ | -0.2284 | V165I Empty Vector | ** (p=0.0051) |
| 5B | WT/WT | -0.1194 | | |
| 5B | WT/W69C | -0.2986 | WT/WT | *** (p=0.0003) |
| 5B | WT/N95S | -0.3346 | WT/WT | *** (p<0.0001) |
| 5B | WT/V165I | -0.3188 | WT/WT | ** (p=0.0012) |

**Supplemental Table 1. Slopes and exact p-values of spotting assay quantifications reported in this manuscript.** The figure reference, test group, slopes, comparison group, and exact p-values are included. ns = not significant.
